# Predictive efficacies of vaccine dose fractionation using neutralizing antibody levels

**DOI:** 10.1101/2022.11.21.22282613

**Authors:** Zhanwei Du, Caifen Liu, Yuan Bai, Lin Wang, Wey Wen Lim, Eric H. Y. Lau, Benjamin J. Cowling

**Affiliations:** WHO Collaborating Centre for Infectious Disease Epidemiology and Control, School of Public Health, Li Ka Shing Faculty of Medicine, The University of Hong Kong, Hong Kong Special Administrative Region, China; Laboratory of Data Discovery for Health Limited (D24H), Hong Kong Science and Technology Park, Hong Kong Special Administrative Region, China; Department of Genetics, University of Cambridge, Cambridge CB2 3EH, UK

**Keywords:** COVID-19, SARS-CoV-2, dose fractionation, neutralizing antibody level

## Abstract

**Background:** With the emergence of SARS-CoV-2 variants that eluded immunity from vaccines and prior infections, vaccine shortages and their effectiveness pose unprecedented challenges for governments to expand booster vaccination programs. Fractionation of vaccine doses might be an effective strategy to help society to face these challenges, which may have comparable efficacies in contrast with the standard doses.

**Methods:** In this study, we analyzed the relationship between in-vitro neutralization levels and the observed efficacies against asymptomatic and symptomatic infection of ten types of COVID-19 vaccines using data from 13 studies from vaccination and convalescent cohorts. We further projected efficacies for fractional doses based on 51 studies included in our systematic review.

**Results:** By comparing with the convalescent level, vaccine efficacy increases from 8.8% (95% CI: 1.4%, 16.1%) to 71.8% (95% CI: 63.0%, 80.7%) against asymptomatic infection, and from 33.6% (95% CI: 23.6%, 43.6%) to 98.6% (95% CI: 97.6%, 99.7%) against symptomatic infection, respectively, along with the mean neutralization level from 0.1 to 10 folds of convalescent level. And mRNA vaccines provide the strongest protection, and decrease slowly for fractional dosing between 50% and 100% dosage.

**Conclusions:** Our results are consistent with studies for immune protection from COVID-19 infection. Based on our study, we expect that fractional dose vaccination could provide a partial immunity for SARS-CoV-2 virus. Fractional doses of vaccines could be a viable vaccination strategy compared to full-dose vaccination and deserves further exploration.

**Key points:** We analyzed the relationship between neutralization levels and efficacies against asymptomatic and symptomatic infection of ten types of COVID-19 vaccines from convalescent cohorts. Fractional doses of vaccines could be a viable strategy compared to full-dose vaccination and deserves further exploration.

---

Coronavirus disease 2019 (COVID-19) continues to threaten fragile healthcare and socioeconomic systems in 2022, exacting a devastating human and economic toll around the world. The primary means of COVID-19 control are the widespread implementation of vaccination and the preservation of public health and social measures. Worldwide, 123 vaccine candidates have been tested in humans by August 31, 2022, with 52 in the final phases of clinical trials [1]. Although 68% of the world population had received at least one dose of the COVID-19 vaccine as of October 1, 2022 [2], global vaccine shortages and inequities persist [3], with only 18.55% and 1.27% of people in low-income countries having been fully vaccinated or receiving booster doses [2].

As of October 2022, the Omicron subvariant BA.5 has displaced BA.2 as the predominant strain of SARS-CoV-2 in countries around the world. Although vaccine effectiveness is expected to wane quickly, a booster shot can restore the protection against infection by both Omicron subvariants to 30–60%, and increase protection against severe disease from a high level to a very high level [4]. A highly vaccinated population is still not enough to combat Omicron’s spread due to immune escape and the waning of vaccine-derived immunity against the ancestral SARS-CoV-2 strain[5]. Countries should time the ramp-up of their booster doses, to account for risks of waning vaccine effectiveness against infection, disease and death.

Given accelerating vaccination and expanding booster programs and concerns about vaccine safety, fractional dosing of vaccines has partial protection to a more significant number of people [6]. For the clinical trial of the mRNA-1273 vaccine, two fractional doses (1/2 of full dose) gave the comparable geometric mean PRNT_80_ titers after two weeks, in contrast with that of two standard doses [7]. Further, the model-predicted efficacy of the half dose and the measured efficacy of the standard dose is all nearly 95% for symptomatic disease [8]. Fractional dose vaccination has successfully addressed vaccine shortages in outbreak events [9]. For example, Angola and the Democratic Republic of Congo adopted fractional doses (1/5 of the standard dose) of the 17DD yellow fever vaccine to accelerate vaccine roll-out during their 2016 yellow fever outbreaks and finally won the war against yellow fever [10,11]. During the monkeypox outbreaks in 2022, the U.S. Food and Drug Administration (FDA) issued an emergency use authorization on August 9, 2022 for the JYNNEOS vaccine to prevent monkeypox infections by intradermal injection in a lower volume (one fifth) to adults 18 years of age and older [12]. During the COVID-19 period, the fractional dose for young people has been approved in some countries considering young adults have a robust immune response to vaccines, and the lower doses may also be linked with fewer side effects. For example, FDA has approved the use of 1/3 of the standard doses of Pfizer (10 μg) for 5- to 11-yr-olds, and 1/10 doses (3 μg) are used in children below 5 years of age [13–15].

Fractional dose vaccination could be an effective strategy for mitigating both epidemic risks by deploying vaccines to reach more individuals in the setting of limited healthcare budgets [8,16], and reduce the disease burden of COVID-19 as compared to the full-dose strategy [17,18]. However, the vaccine efficacy has been tested mainly for standard doses by clinical trials [19], rather than fractional doses which are measured by phase 1 and 2 clinical trials of various vaccines with immune response in the form of neutralizing antibody (NAb) titers. Ref. [8] derived the efficacy against symptomatic infection of fractional doses using its relationship with NAb titers in standard doses [19]. However, we are still unclear about other key efficacy (e.g., a/symptomatic infection) for standard and fractional doses. Here we investigate the relationship among NAb titers and the vaccine efficacies against disease for standard doses and derive vaccine efficacies for fractional doses, informed by real-world data through conducting a systematic review of vaccination efficacy for both standard and fractional doses.

## METHODS

### Data source and searches

We performed a systematic review of peer-reviewed studies on within-host models of SARS-CoV-2 in PubMed on 15 March 2022. We searched studies in PubMed with a combination of the following search terms, with no restriction on publication language: (#1) “COVID-19” OR “SARS-CoV-2” OR “2019-nCoV” OR “coronavirus”; (#2) “vaccin*”; (#3) “fractiona*” OR “dose”; (#4) “efficacy” OR “effectiveness” OR “neutralizing antibody” OR “neutralising antibody” OR “neutralization titer” OR “neutralization level” OR “antibody titer” OR “immune response” OR “immune protection” OR “immunogenicity” OR “reactogenicity” OR “safety” OR “adverse event” OR “adverse reaction” OR “adverse effect”; and the final search term was #1 AND #2 AND #3 AND #4. The searched studies were set to be published between 1 January 2020 and 15 March, 2022.

### Study selection

We (Caifen Liu and Yuan Bai) assessed eligible studies, extracted relevant data, and conducted cross-checked. Conflicts over the study selection were resolved by another researcher (Zhanwei Du). We excluded studies based on screening titles and abstracts if they were 1) reviews, commentaries, preprints; 2) not about SARS-CoV-2 vaccines; 3) preliminary animal studies; 4) focusing on full-dose vaccines; or 5) not about neutralizing antibody response. We excluded studies based on full-text assessment if they were 1) lack of original neutralizing antibody data; 2) reporting vaccines that were no longer processing; or 3) reporting vaccines whose standard dosage were not determined. We reported studies following the Preferred Reporting Items for Systematic Reviews and Meta-Analyses (PRISMA) guidelines.

### Data extraction and analysis

Information relevant to vaccines and participants was extracted from the selected studies, which include vaccine name, platform, standard dosage (defined as the dosage determined for the approved vaccine or phase 3 trials), vaccination schedule, neutralization assay, neutralization measured date, target virus/pseudovirus used to test NAb response, sample size and age group of vaccinated participants, and sample size of convalescents (**Table S1**). For each study, the mean neutalizing titers coupled with the 95% confidence intervals (CIs) were extracted. Studies not reporting neutralizing titers of convalescents were excluded for further analysis. A random-effects model or a fixed effect model was further used to perform a meta-analysis according to the assessed heterogeneity between studies. Analyses were conducted in R version 4.1.1.

## Statistical methods

### Mean neutralization level

To compare neutralization titers across studies using different assays, we standardized the NAbs level as the ratio of the vaccine-induced NAbs level versus NAbs measured in convalescent sera for each study [19], and refer to this ratio as the ‘mean neutralization level’ in our study. Note that the log of the mean neutralization level for conconvalescents is equal to zero by definition. Following the assumption by Ref. [19], the log_10_ transform of the mean neutralization level is normally distributed. For dose group ***j*** in study ***s***, we calculated the the ratio of mean NAbs titers of vaccine group versus mean NAbs titers of convalesents group to estimate the corresponding log_10_ transformed mean neutralization level ***μ***_***sj***_ of this dose group, and used standard deviation of vaccine group to estimate ***σ***_***sj***_. We used bootstrap method based on the normal distribution with estimated mean 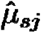 and standard deviation 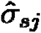 from 2000 bootstrap samples to obtain the 95% confidence intervals as 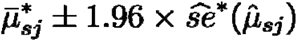 and 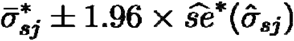, where 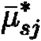 and 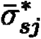 are the mean of bootstrap replicates for 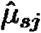 and 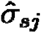, respectively. For each fractional dosing in a study, we extracted reported NAbs measured at different time points and calculated their mean neutralization levels accordingly (**Figure S3**). Over the study period after full vaccination for each dosage, the peak level of NAbs was selected to model the relationship between NAbs level and vaccine efficacy.

### Logistic method

Informed by the mean neutralization levels and their efficacies from each vaccine in eligible studies, we fitted the logistic model to study the relationship between neutralization and protection against infection outcomes (***o***, i.e., asymptomatic or symptomatic infection), following Ref. [19] (**Figure S4**):

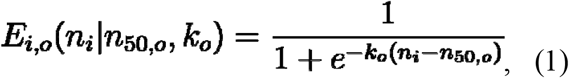

where ***E***_***i***,***o***_ is the protective efficacy of an individual ***i*** against infection outcome ***o*** given the neutralization level ***n***_***i***_, and ***n***_***i***_ is the log_10_ transform of the mean neutralization level. The paramater ***n***_**50, *o***_ denotes the log_10_-transformed mean neutralization level at which an individual will have 50% efficacy against infection outcome ***o***, and ***k***_***o***_ is the corresponding steepness of this model. We assumed the vaccine-induced and infection-induced ***n***_***i***_ follows a normal distribution with mean ***μ***_***s***_ and standard deviation ***σ***_***s***_ in study ***s***. The proportion of the vaccinated population that will be protected is given by:

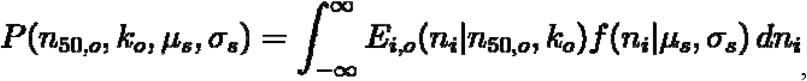

where ***f*** is the normal probability density function of ***n***_***i***_. The mean parameter ***μ***_***s***_ was estimated by the mean neutralization level of each study and the estimated ***σ***_***s***_ was calculated as 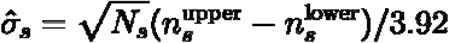, where 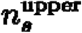 and 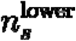 denote the upper and lower bound of the 95% CI of ***n***_***s***_, respectively, and ***N***_***s***_ is the sample size of the phase 3 in study ***s***.

Based on the efficacy data reported from phase 3 clinical trials, along with the number of enrolled participants of placebo and vaccine group and infections in each group (i.e., the number of participants in placebo group 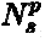, the number of participants in vaccine group 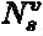, the number of infected participants in placebo group 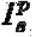, the number of infected participants in vaccine group 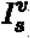), we obtained the likelihood of the observed infections of outcome ***o*** in study ***s***:

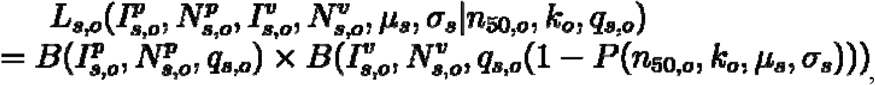

where ***B(K, N, q)***denotes the binomial probability mass function of the probability of getting ***K*** events from a sample size of ***N*** with a success probability ***q, q***_***s, o***_. is the probability of an unvaccinated individual to develop infection outcome ***o*** in study ***s***.

***q*** _***s***,***o***_ **(1 −*P*(*n***_**50, *o***_, ***k***_***o***_, ***μ***_***s***_, ***σ***_***s***_))is that probability in the vaccine group accordingly. The likelihood of observing infections of outcome ***o*** for all studies is the product of the likelihood for each study:

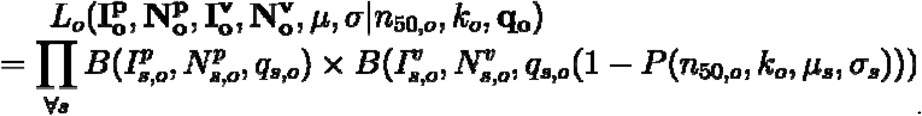

The maximum likelihood estimates of ***n***_**50**,***o***,_ ***k***_***o***_ and **q**_**o**_ were obtained using *nlm* function in R (programming language) by minimizing -log (*L*_*o*_). The standard error of the estimates was computed using the output Hessian matrix ***H*** as 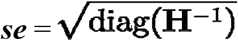, and the 95% CIs were calculated as 1.96 *se* of the estimates.

### Projection of vaccine efficacies

Using the fitted logistic model, we projected the vaccine efficacy against asymptomatic and symptomatic infection over fractional dosing for each vaccine type. We categorized different doses into four groups, i.e., dose <50%, dose = 50%, 50% < dose < 100%, and dose = 200% (the fraction was calculated as the ratio of each tested dose group versus the standard dose). For each vaccine type included in the final analysis, namely, mRNA, protein subunit, non-replicating viral vector, inactivated, and DNA vaccine, we pooled the mean neutralization levels by dose group and predicted the efficacy of each dose group using the pooled mean. By substituting the pooled mean neutralization level for ***n***_***i***_ into Eq. (1), we obtained the predicted vaccine efficacy for each fractional dosing (**Figure 1**).

**Figure 1.**
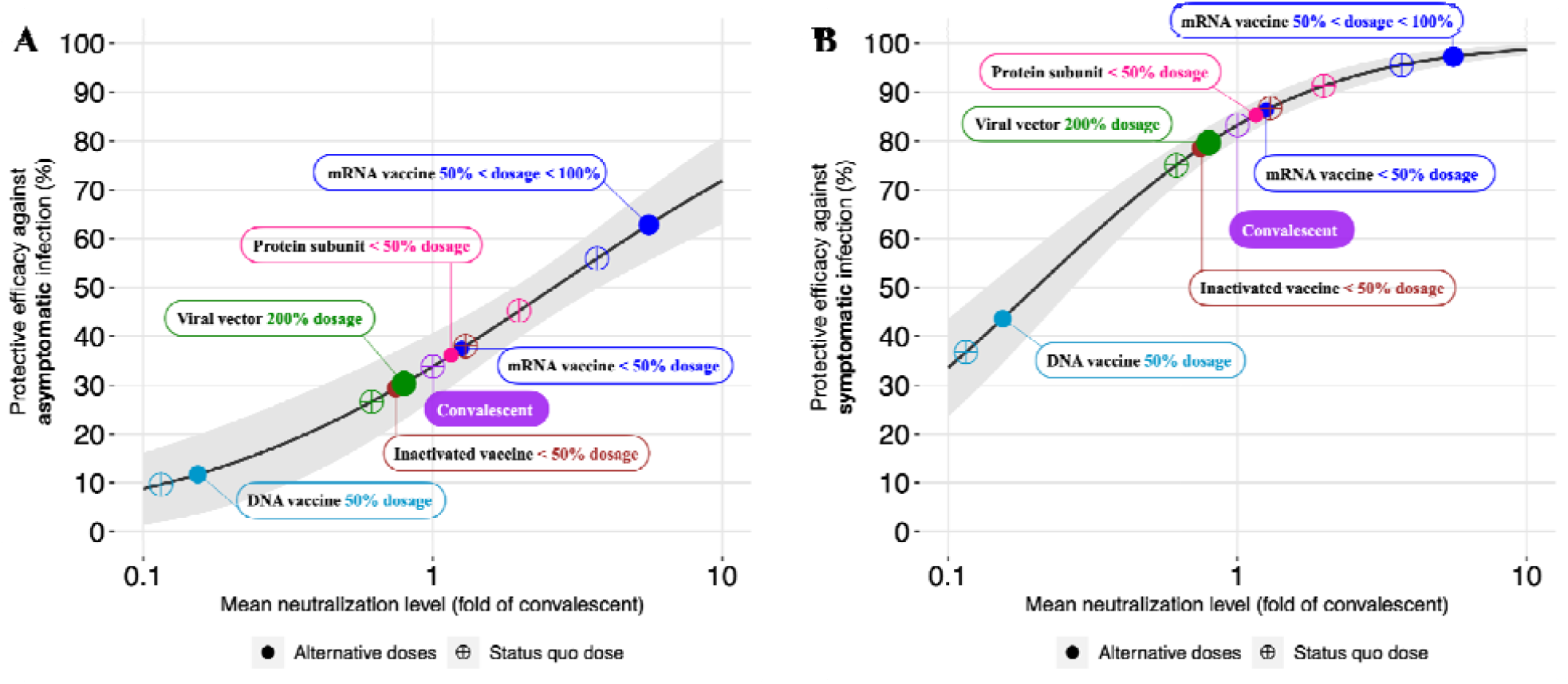
Estimated vaccine efficacy using neutralizing antibody levels. Estimated vaccine efficacy against (**A**) asymptomatic infection, and (B) symptomatic infection. The x-axis and y-axis denote the reported mean neutralization level from phase 1 and 2 trials and the predicted protective efficacy for vaccines or the convalescent cohort, respectively. The gray solid line and shading indicate the best fit and the 95% confidence interval of the logistic model. Each dot indicates values of predicted vaccine efficacy of vaccines (e.g., mRNA, Protein subunit, viral vector, inactivated, and DNA) or convalescent individuals.

To illustrate the dose-efficacy relationship for different vaccines, we first quantified the relationship between dose fraction and mean neutralization levels using a generalized additive model such that:

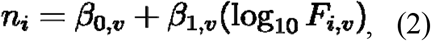

where ***n***_***i***_ is the log_10_-transformed mean neutralization level of individual ***i, F*** _***i***,***v***_ is the dose fraction of vaccine type ***v*** (**Figure 2**). A non-parametric bootstrap method was conducted to obtain the estimated standard error of 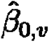 and 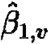 based on 2000 replicates. Combining Eq. (2) with Eq. (1), the dose-efficacy relationship was derived (**Figure 3-6**).

**Figure 2.**
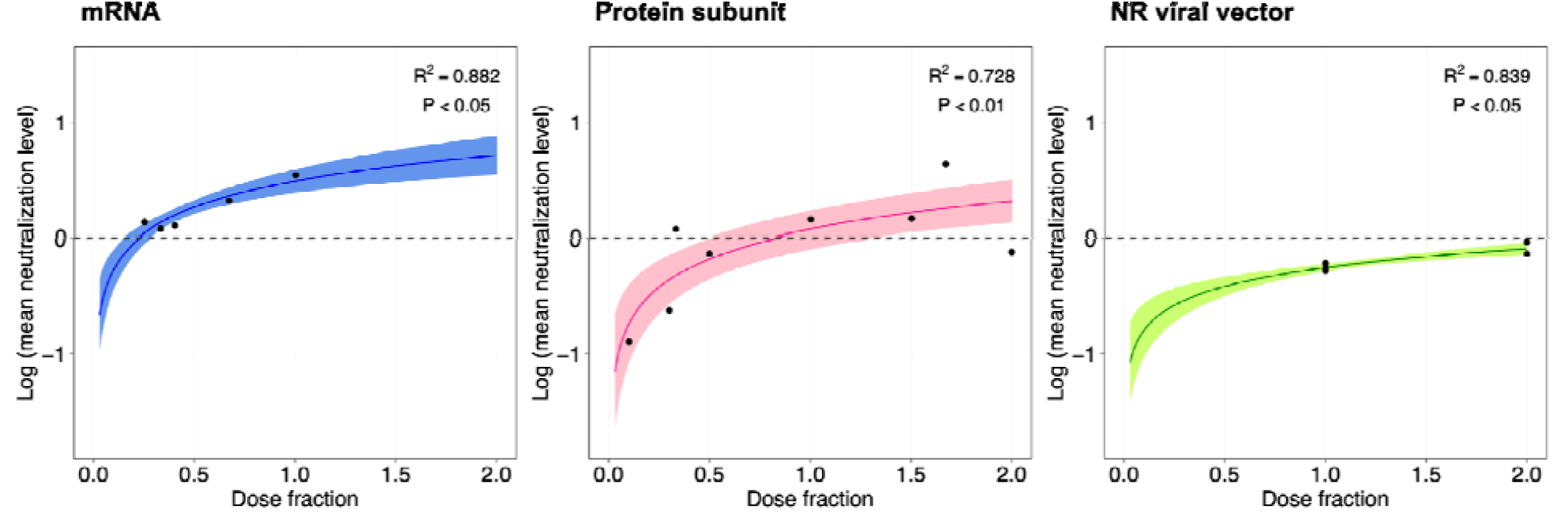
Model fit of neutralizing antibody levels using dose fraction. The x-axis and y-axis denote the dose fraction and the estimated log_10_-transformed neutralizing antibody level for three types of vaccines (e.g., mRNA, Protein subunit, and Non-replicating viral vector), respectively. The solid line and shading indicate the best fit and the 95% confidence interval of the logistic model.

**Figure 3.**
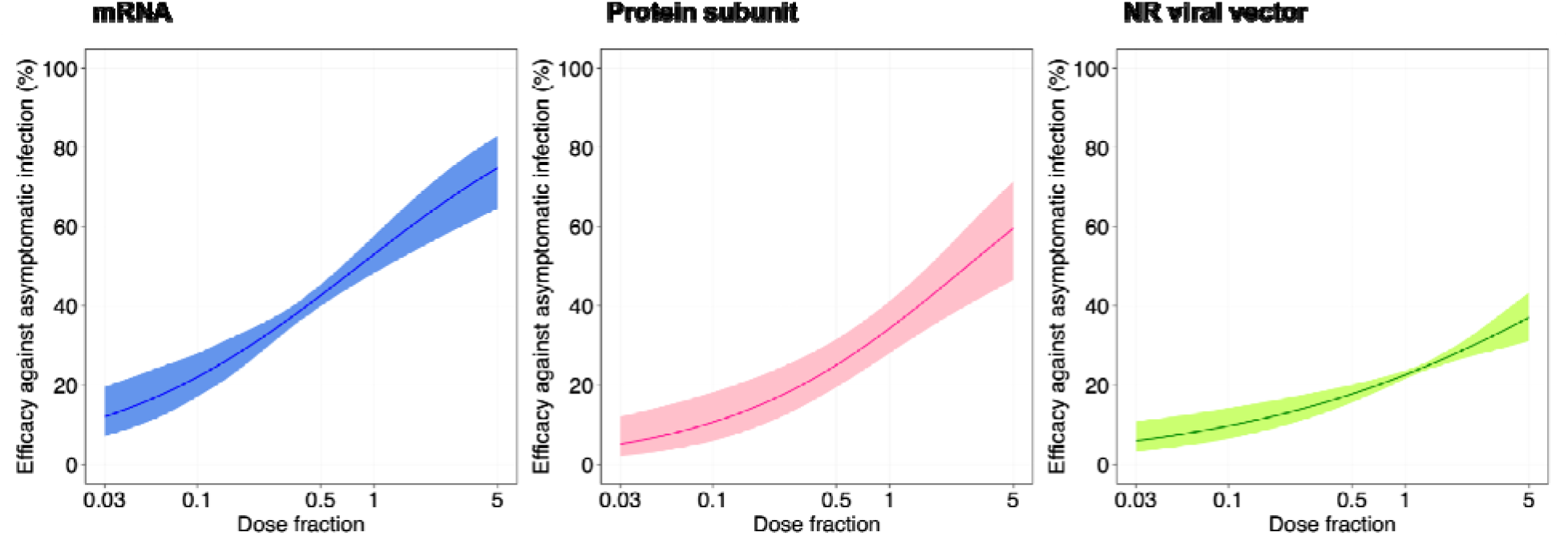
Estimated vaccine efficacy against asymptomatic infection using neutralizing antibody levels over vaccines. The x-axis and y-axis denote the fraction of dose and the estimated protective efficacy for three types of vaccines (e.g., mRNA, Protein subunit, and Non-replicating viral vector), respectively. The gray solid line and shading indicate the best fit and the 95% confidence interval of the logistic model.

## RESULTS

We identified 2811 studies through the electronic search on PubMed between 1 January 2020 and 15 March, 2022. 2777 studies left after excluding duplicates. After 2694 studies were excluded based on titles and abstracts screening, we retrieved 83 studies eligible for the full-text screening. Next, after we excluded 32 studies based on full-text screening, 51 studies met the inclusion criteria and were included in the systematic review (**Table S1, Figure S1, Figure S2**).

Alongside these published studies included in systematic review, we collected 13 records of vaccine efficacy against asymptomatic and symptomatic infection for 11 COVID-19 variants (e.g., Alpha, Beta, and Delta) of 10 types of vaccines (e.g., BNT162b2, ChAdOx1 nCov-19, and CoronaVac) from Pubmed (**Table S2**). Given various assays used in each study, we projected neutralization titers to the mean convalescent titer using the same assay following the method in Ref. [19]. Informed by the information of standard-dose vaccines, the non-linear relationship between the standardized mean neutralization level and each of the four reported vaccine efficacies is fitted by the logistic model (**Figure S4**), which provides a good explanation of the study relationship (rooted mean square error (RMSE) for predicted vaccine efficacy is 2.43 and 0.16 [%] for asymptomatic and symptomatic infection, respectively). We predicted the vaccine efficacy against infection outcomes of fractional doses based on the established model (**Figure 1**). By comparing with the convalescent level, vaccine efficacy against asymptomatic and symptomatic infection increases from 8.8% (95% CI: 1.4%, 16.1%) to 71.8% (95% CI: 63.0%, 80.7%), and from 33.6% (95% CI: 23.6%, 43.6%) to 98.6% (95% CI: 97.6%, 99.7%), respectively, along with the mean neutralization level from 0.1 to 10 folds of convalescent level (**Figure 1**). And mRNA vaccines provide the strongest protection, and decrease slowly for fractional dosing between 50% and 100% dosage.

We further estimated the potential levels of protection that could be provided by fractional dosing for each type of vaccine. mRNA and Protein subunit vaccines have a significantly increasing trend of vaccine efficacies over dose fraction (**Figure 2, Figure 3, Figure 4**). For example, the mRNA vaccine efficacy against asymptomatic and symptomatic infection increase from 12.1% (95% CI: 7.2%, 19.7%) to 74.8% (95% CI: 64.6%, 82.9%), and 52.9% (95% CI: 29.8%, 74.8%) to 99.5% (95% CI: 98.9%, 99.8%), respectively, from 0.03 dosing to 5 dosing. The Protein subunit vaccines have a efficacy of 5.2% (95% CI: 2.1%, 12.2%) to 59.6% (95% CI: 46.5%, 71.4%) against asymptomatic infection, and 19.4% (95% CI: 4.8%, 53.2%) to 98.4% (95% CI: 96.2%, 99.3%) against symptomatic infection in the study range of dose fractionation from 0.03 to 5. The non-replicating viral vector vaccines have a efficacy of 6.0% (95% CI: 3.3%, 10.9%) to 37.1% (95% CI: 31.2%, 43.4%) against asymptomatic infection, and 23.6% (95% CI: 9.5%, 47.7%) to 92.9% (95% CI: 89.3%, 95.3%) against symptomatic infection in the study range of dose fractionation from 0.03 to 5.

**Figure 4.**
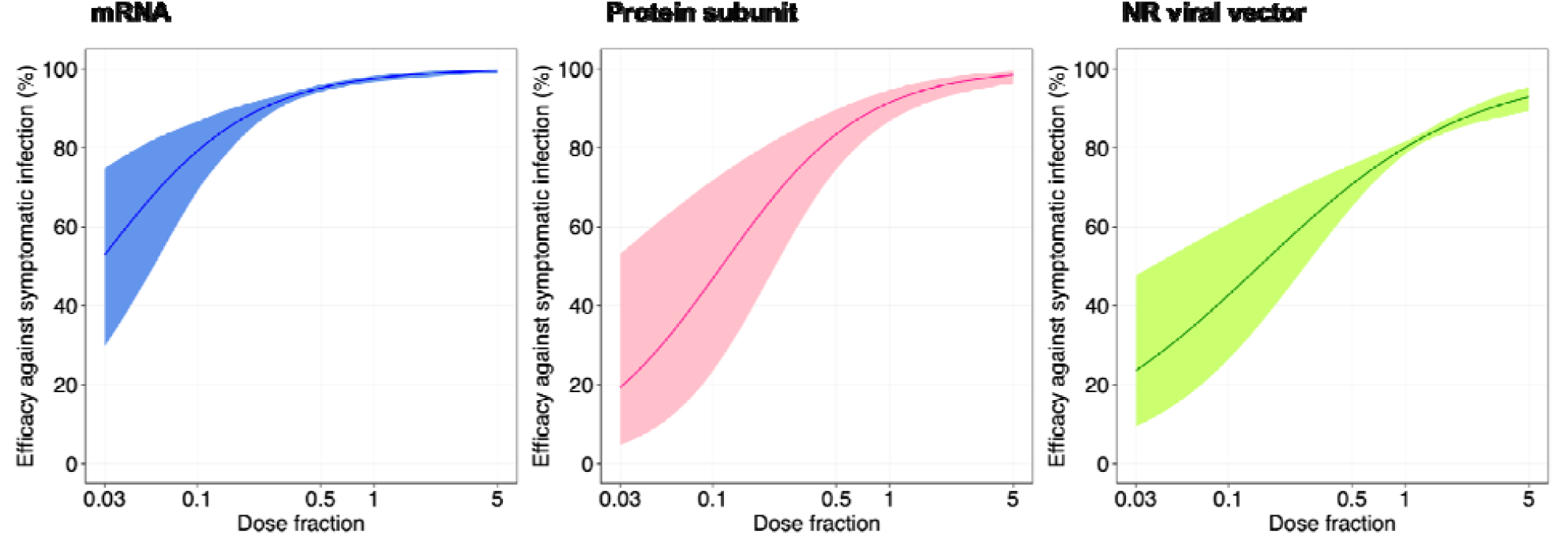
Estimated vaccine efficacy against symptomatic infection using neutralizing antibody levels over vaccines. The x-axis and y-axis denote the fraction of dose and the estimated protective efficacy for three types of vaccines (e.g., mRNA, Protein subunit, and Non-replicating viral vector), respectively. The solid line and shading indicate the best fit and the 95% confidence interval of the logistic model.

## DISCUSSION

The immunogenicity, efficacy, and safety of fractional doses have been reviewed as of December 2021, indicating that they would be safe and effective comparable to the standard dose [20]. We further investigate the relationship among NAb titers and the vaccine efficacies against infection for standard doses and derive vaccine efficacies for fractional doses, informed by available data from published studies of vaccination efficacy for both standard and fractional doses. It suggests efficacies are predictive using neutralizing antibody levels for both standard- and fractional-dosing vaccines.

There would be public health and economic advantages to using a dose-sparing strategy to increase vaccine supply and vaccination coverage around the world [8,16]. According to the World Health Organization (WHO), fractional dosing strategies have the potential to save lives, but only after a thorough review of immunogenicity data [21]. Using an untested fractionated vaccine dose in a low-resource setting should also be considered ethical and political, as higher-resource settings have access to full vaccine doses [16]. The fractional dose vaccination could help to optimize the availability of vaccine doses with vaccine shortage. With vaccine supply increasing, this strategy would still have a high value for vulnerable individuals with limited side effects.

In spite of the robustness of our qualitative results, we have identified some limitations. First, our study does not explicitly include age groups, which may have different levels of vaccine efficacy and safety. Second, our study does not include the waning rate of vaccine-derived immunity for fractional dose vaccination, which may vary with respect to dose size [8]. Third, the NAbs data are all ancestral stains, but efficacy data consists of different variants. Ideally, we should use NAbs and efficacy data of the same strain, but NAbs data of different variants were limited, which may have different levels of protection than that against VOCs and for booster doses. For example of ChAdOx1 (Covishield, AstraZeneca) vaccine, effectiveness against symptomatic disease is 17.7%, 33.7%, and 60% for the Omicron variant in the fifth week post the first, second and booster doses, respectively, but higher levels of 42.9%, 76.5%, and 94.9% for the Delta variant [22]. Fourth, the included clinical studies target 12 COVID-19 variants (e.g., wild-type, Alpha, Beta, and Delta). The same vaccine may have different protection performance for different variants. However, this will not impact the relationship between neutralizing antibody levels and vaccine efficacies.

To summarize, our results are consistent with studies for predictive immune protection from symptomatic COVID-19 infection [19] and we further projected vaccine protections against asymptomatic. We expect that fractional dose vaccination could provide a partial immunity (e.g., against infection) for SARS-CoV-2 virus. This strategy would be cost-effective to curb coronavirus disease outbreak by balancing the lower efficacy, faster vaccination coverage, and lower side effects, especially if global vaccine shortages issues persist.

## Data Availability

All data are collected from open source with a detailed description in the Methods section.

## Notes

## Acknowledgments

We acknowledge the financial support from the AIR@InnoHK Programme from Innovation and Technology Commission of the Government of the Hong Kong Special Administrative Region, and the National Institute of Allergy and Infectious Diseases, National Institutes of Health, Department of Health and Human Services, under contract no. 75N93021C00015. The funders of the study had no role in study design, data collection, data analysis, data interpretation, or writing of the report. We thank the research assistants of Shuqi Wang, Zengyang Shao, and Mingda Xu for their help to review and check eligible studies.

## Author Contributions

ZD, CL, and BJC: conceived the study, designed statistical and modeling methods, conducted analyses, interpreted results, wrote and revised the manuscript; YB and EHYL: interpreted results and revised the manuscript.

## Competing interests

BJC reports honoraria from AstraZeneca, Fosun Pharma, GlaxoSmithKline, Moderna, Pfizer, Sanofi Pasteur, and Roche. The authors report no other potential conflicts of interest.

## Data availability

All data are collected from open source with a detailed description in the Methods section.

## Code availability

The code used for data analysis is freely available upon request.

## References

1. Corum J, Wee S-L, Zimmer C. Coronavirus Vaccine Tracker. The New York Times. 2020; Available at: https://www.nytimes.com/interactive/2020/science/coronavirus-vaccine-tracker.html. Accessed 25 September 2020.

2. Ritchie H, Mathieu E, Rodés-Guirao L, et al. Coronavirus Pandemic (COVID-19). Our World in Data 2020; Available at: https://ourworldindata.org/covid-vaccinations. Accessed 14 April 2022.

3. Usher AD. A beautiful idea: how COVAX has fallen short. Lancet 2021; 397:2322–2325.

4. Sidik SM. Vaccines protect against infection from Omicron subvariant - but not for long. Nature 2022; Available at: http://dx.doi.org/10.1038/d41586-022-00775-3.

5. Mallapaty S. China’s zero-COVID strategy: what happens next? Nature 2022; 602:15–16.

6. Cowling BJ, Lim WW, Cobey S. Fractionation of COVID-19 vaccine doses could extend limited supplies and reduce mortality. Nat Med 2021; 27:1321–1323.

7. Jackson LA, Anderson EJ, Rouphael NG, et al. An mRNA Vaccine against SARS-CoV-2 — Preliminary Report. N Engl J Med 2020; 383:1920–1931.

8. Więcek W, Ahuja A, Chaudhuri E, et al. Testing fractional doses of COVID-19 vaccines. Proc Natl Acad Sci U S A 2022; 119. Available at: http://dx.doi.org/10.1073/pnas.2116932119.

9. Ali A, Jafri RZ, Messonnier N, et al. Global practices of meningococcal vaccine use and impact on invasive disease. Pathog Glob Health 2014; 108:11–20.

10. Casey RM, Harris JB, Ahuka-Mundeke S, et al. Immunogenicity of Fractional-Dose Vaccine during a Yellow Fever Outbreak — Final Report. New England Journal of Medicine. 2019; 381:444–454. Available at: http://dx.doi.org/10.1056/nejmoa1710430.

11. Wu JT, Peak CM, Leung GM, Lipsitch M. Fractional dosing of yellow fever vaccine to extend supply: a modelling study. Lancet 2016; 388:2904–2911.

12. Office of the Commissioner. Monkeypox Update: FDA Authorizes Emergency Use of JYNNEOS Vaccine to Increase Vaccine Supply. FDA, Available at: https://www.fda.gov/news-events/press-announcements/monkeypox-update-fda-authorizes-emergency-use-jynneos-vaccine-increase-vaccine-supply. Accessed 12 October 2022.

13. Pitts J, Triano C, Alatovic J, Maas S. Pfizer-BioNTech Announce Positive Topline Results of Pivotal COVID-19 Vaccine Study in Adolescents. 2021; Available at: https://www.pfizer.com/news/press-release/press-release-detail/pfizer-biontech-announce-positive-topline-results-pivotal.

14. Office of the Commissioner. FDA authorizes Pfizer-BioNTech COVID-19 Vaccine for emergency use in children 5 through 11 years of age. 2021. Available at: https://www.fda.gov/news-events/press-announcements/fda-authorizes-pfizer-biontech-covid-19-vaccine-emergency-use-children-5-through-11-years-age. Accessed 24 March 2022.

15. Office of the Commissioner. Coronavirus (COVID-19) Update: FDA Authorizes Moderna and Pfizer-BioNTech COVID-19 Vaccines for Children Down to 6 Months of Age. FDA, Available at: https://www.fda.gov/news-events/press-announcements/coronavirus-covid-19-update-fda-authorizes-moderna-and-pfizer-biontech-covid-19-vaccines-children. Accessed 4 October 2022.

16. Du Z, Wang L, Pandey A, et al. Modeling comparative cost-effectiveness of SARS-CoV-2 vaccine dose fractionation in India. Nat Med 2022; 28:934–938.

17. Tuite AR, Zhu L, Fisman DN, Salomon JA. Alternative Dose Allocation Strategies to Increase Benefits From Constrained COVID-19 Vaccine Supply. Ann Intern Med 2021; 174:570–572.

18. Barnabas RV, Wald A. A Public Health COVID-19 Vaccination Strategy to Maximize the Health Gains for Every Single Vaccine Dose. Ann Intern Med 2021; 174:552–553.

19. Khoury DS, Cromer D, Reynaldi A, et al. Neutralizing antibody levels are highly predictive of immune protection from symptomatic SARS-CoV-2 infection. Nat Med 2021; Available at: http://dx.doi.org/10.1038/s41591-021-01377-8.

20. Yang B, Huang X, Gao H, Leung N, Tsang T, Cowling B. Immunogenicity, efficacy, and safety of SARS-CoV-2 vaccine dose fractionation: a systematic review and meta-analysis. Research Square. 2022; Available at: https://www.researchsquare.com/article/rs-1571821/latest.pdf.

21. Interim statement on dose-sparing strategies for COVID-19 vaccines (fractionated vaccine doses). Available at: https://www.who.int/news/item/10-08-2021-interim-statement-on-dose-sparing-strategies-for-covid-19-vaccines-(fractionated-vaccine-doses). Accessed 11 December 2021.

22. Andrews N, Stowe J, Kirsebom F, et al. Covid-19 Vaccine Effectiveness against the Omicron (B.1.1.529) Variant. N Engl J Med 2022; Available at: http://dx.doi.org/10.1056/NEJMoa2119451.

